# A short report: Acute, non-COVID related medical admissions during the first wave of COVID-19: A retrospective comparison of changing patterns of disease

**DOI:** 10.1101/2020.09.15.20194795

**Authors:** B Riley, M Packer, S Gallier, E Sapey, C Atkin

## Abstract

**Background:** The COVID-19 pandemic was associated with social restrictions in the UK from 16^th^ March 2020. It was unclear if the lockdown period was associated with differences in the case-mix of non-COVID acute medical admissions compared with the previous year.

**Methods:** Retrospective data were collected for 1^st^-30^th^ April 2019 and 1^st^–30^th^ April 2020 from University Hospitals Birmingham NHS Foundation Trust, one of the largest hospitals in the UK with over 2 million patient contacts per year. The latter time period was chosen to coincide with the peak of COVID-19 cases in the West Midlands. All patients admitted under acute medicine during these time periods were included. COVID-19 was confirmed by SARS-Cov-2 swab or a probable case of COVID-19 based on World Health Organization diagnostic parameters. Non-COVID patients were those with a negative SARS-Cov-2 swab and no suspicion of COVID-19. Data was sourced from UHB’s in-house electronic health system (EHS).

**Results:** The total number of acute medical admissions fell comparing April 2019 (n = 2409) to April 2020 (n = 1682). As a proportion of total admissions, those aged under 45 years decreased, while those aged 46 and over did not change.

The number of admissions due to psychiatric conditions and overdoses was higher in April 2020 (p < 0.001). When viewed as a proportion of admissions, alcohol-related admissions (p = 0.004), psychiatric conditions and overdoses (p< 0.001) increased in April 2020 than in April 2019. The proportion of patients who were in hospital due to falls also increased in April 2020 (p< 0.001). In the same period, the absolute number and the proportion of admissions that were due to non-specific chest pain, to musculoskeletal complaints and patients who self-discharged prior to assessment decreased (p = 0.02, p = 0.01 and p = 0.002 respectively).

There were no significant differences in non-COVID-related intensive care admissions or mortality between the same months in the two years.

**Conclusion:** In this large, single-centre study, there was a change in hospitalised case-mix when comparing April 2019 with April 2020: an increase in conditions which potentially reflect social isolation (falls, drug and alcohol misuse and psychiatric illness) and a decrease in conditions which rarely require in-patient hospital treatment (musculoskeletal pain and non-cardiac chest pain) especially among younger adults. These results highlight two areas for further research; the impact of social isolation on health and whether younger adults could be offered alternative health services to avoid potentially unnecessary hospital assessment.

## Introduction

On 12^th^ March 2020, the World Health Organization declared the novel coronavirus SARS-CoV-2 as a pandemic. The UK government strategy to mitigate risk led to a national lockdown (implemented 23/3/20). There was dramatic and rapid restructuring of acute NHS services in both primary and secondary care to meet the unprecedented change in clinical need.

The majority of unplanned admissions to acute hospitals are medical emergencies, with patients assessed and treated by the acute medical team^(1)^. Anecdotally, during the peak of the pandemic there appeared to be a decline in patient numbers admitted to medicine for reasons other than COVID-19. This corresponded to a reduction in the overall number of attendances to emergency departments nationally^(2)^. A decrease in hospital admissions for specific diagnoses, such as acute coronary syndrome, has been demonstrated^(3)^however the impact of the COVID-19 pandemic on medical admissions overall has not been explored.

The aim of this study was to compare medical admissions during the height of the first wave of the COVID-19 pandemic with an equivalent pre-pandemic period in a large secondary care hospital which has seen a significant burden of COVID-19 cases and deaths^(4).^

## Methods

University Hospitals Birmingham NHS Foundation Trust (UHB), UK is one of the largest Trusts nationally, treating over 2.2 million patients per year and housing the largest single critical care unit (CCU) in Europe. UHB saw the highest number of COVID admissions in the UK (3670 confirmed cases by 27th August 2020) and the highest number of patients ventilated, with an expanded CCU capacity of > 200 beds.

Retrospective data was collected for 1^st^-30^th^ April 2019 and 1^st^–30^th^ April 2020. This time period coincided with the peak of COVID-19 cases in the West Midlands^(4)^. All patients admitted under acute medicine during these time periods were included. Patients referred through the acute stroke or ST elevation myocardial infarctions pathway, direct specialty admissions including from the emergency department to CCU were not included. COVID-19 was confirmed by SARS-Cov-2 swab or a probable case of COVID-19 based on World Health Organization diagnostic parameters^(5)^. Non-COVID patients were those with a negative SARS-Cov-2 swab and no suspicion of COVID-19.

Data was sourced from UHB’s in-house electronic health system (EHS); Prescribing Information and Communication System (PICS). Data collected included patient demographics, presenting complaint, length of stay, discharge diagnosis, DNACPR status, critical care admission, and death during that admission episode. Discharge diagnoses were taken from electronic discharge documentation and bereavement documentation for patients who died during admission. Diagnoses were grouped into broad categories by clinicians. Any patients without a diagnosis on their discharge summary had their notes reviewed by a clinician and was categorised accordingly.

Statistical analysis was performed using SPSS, version 26 (IBM). Continuous variables were compared between data sets using Mann-Whitney U tests. Categorical variables were compared using Fisher exact and Chi-Square tests. Results were considered significant if the p-value was < 0.05.

This work was supported by PIONEER, a Health Data Research Hub in Acute Care with ethical approval provided by the East Midlands – Derby REC (reference: 20/EM/0158).

## Results

The total number of medical admissions fell comparing April 2019 (n = 2409) to April 2020 (n = 1682). Patient demographics are shown in Table 1. Patients admitted in April 2020 were older than in April 2019 (median age (IQR) 2020: 69 years (52-82), 2019: 66 years (47-80), p< 0.001). Age distribution of admissions is shown in Figure 1. As a proportion of total admissions, those aged 16–45 years decreased, while those aged 46-65 years, 66–75 years and over 76 years did not change.

**Figure 1:**
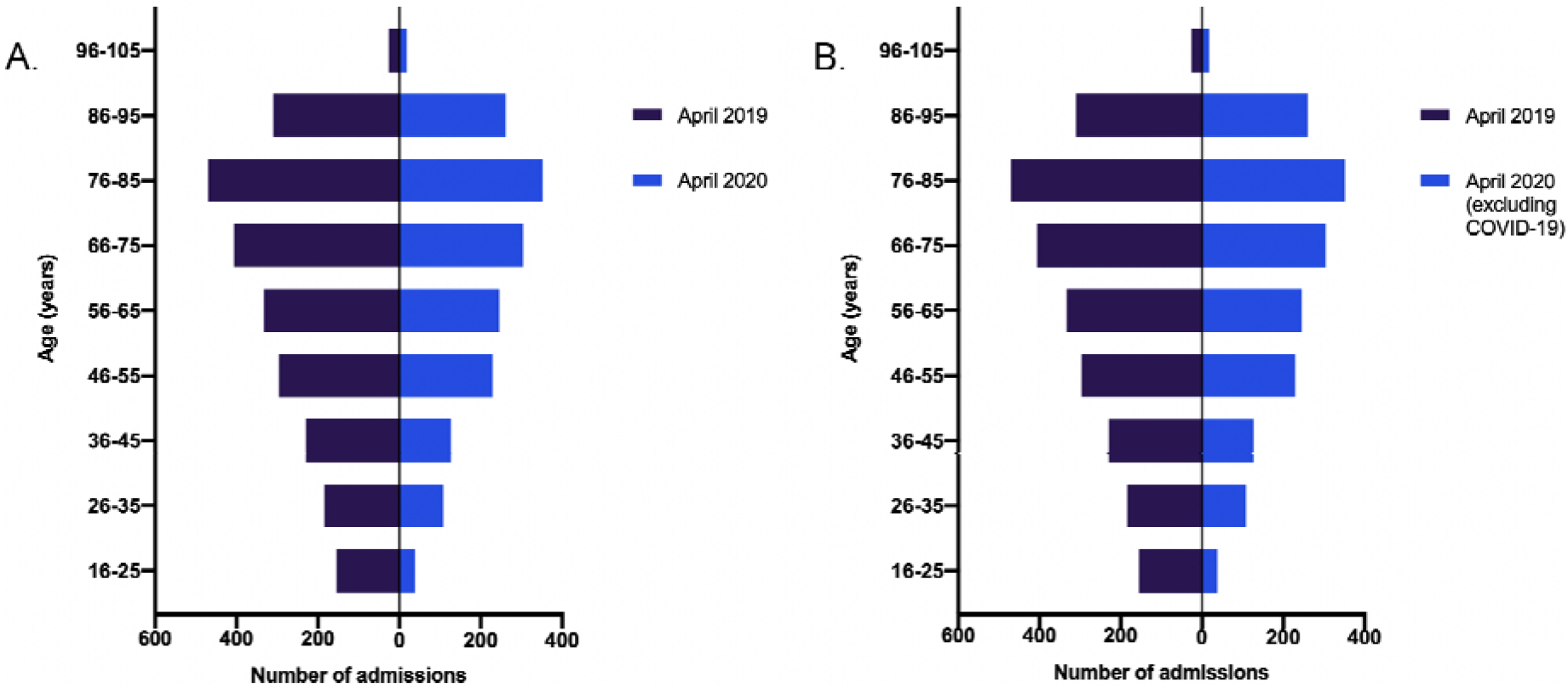
The age distribution of all medical admissions. **Legend:** A. All medical admissions in April 2019 and April 2020 and B. All medical admissions in April 2019 and April 2020 (excluding patients with COVID-19). For both graphs, April 2019 patients are shown in dark blue and April 2020 patients are shown in mid-blue. Patients (both with and excluding COVID-19 presentations) were older in the 2020 cohort.

**Table 1:** Patient characteristics for admissions in April 2019 and April 2020. **Legend.** Diagnosis taken from discharge documentation for patients discharged from hospital, or from bereavement documentation for patients who died during admission. P-values comparing all admissions April 2019 vs all admissions April 2020 and comparing all admissions April 2019 vs Admissions in April 2020 (excluding those with a discharge diagnosis of COVID-19). P values calculated using Mann-Whitney U test for continuous data (age, length of stay), Chi square for categorical variables (sex, critical care admission, mortality, documented DNACPR order). CCU = critical care unit; DNACPR = do not attempt cardiopulmonary resuscitation; IQR = interquartile range; LOS = length of stay; Mortality = in-hospital mortality. For age, p values in bold, comparing the proportion of adults aged 16 – 45 years, then 46 – 65 years, 66 – 75 years and over 76 years presenting in 2019 to 2020 (all admissions) and 2020 (excluding COVID-19).

|  | April 2019 |  | April 2020 |  |  |  |  |  |  |  |
| --- | --- | --- | --- | --- | --- | --- | --- | --- | --- | --- |
|  | All acute medicine admissions |  | All acute medicine admissions |  | p value (to April 2019) | Admissions excluding those due to COVID-19 |  | p value (to April 2019) | Admissions due to COVID-19 |  |
|  | N | % | N | % |  | N | % |  | N | % |
| **Admission** | 2409 |  | 1682 |  |  | 1222 |  |  | 460 |  |
| **Age (yrs)** |  |  |  |  |  |  |  |  |  |  |
| Median (IQR) | 66 (47-80) |  | 69 (52-82) |  | <0.001 | 68 (51-82) |  | 0.03 | 70.5 (54-83) |  |
| 16-25 | 155 | 6.43 | 38 | 2.26 |  | 33 | 2.70 |  | 5 | 1.09 |
| 26-35 | 184 | 7.64 | 108 | 6.42 | <0.0001 | 91 | 7.45 | <0.0001 | 17 | 3.70 |
| 36-45 | 229 | 9.51 | 127 | 7.55 |  | 93 | 7.61 |  | 34 | 7.39 |
| 46-55 | 296 | 12.29 | 229 | 13.61 | 0.142 | 164 | 13.42 | 0.219 | 65 | 14.13 |
| 56-65 | 333 | 13.82 | 245 | 14.57 |  | 179 | 14.65 |  | 66 | 14.35 |
| 66-75 | 406 | 16.85 | 304 | 18.07 | 0.314 | 227 | 18.58 | 0.195 | 77 | 16.74 |
| 76-85 | 470 | 19.51 | 352 | 20.93 | 0.421 | 244 | 19.97 | 0.208 | 108 | 23.48 |
| 86-95 | 310 | 12.87 | 261 | 15.52 |  | 180 | 14.73 |  | 81 | 17.61 |
| 96+ | 26 | 1.08 | 18 | 1.07 |  | 11 | 0.90 |  | 7 | 1.52 |
| **Male Sex** | 1054 | 43.75 | 881 | 52.38 | <0.001 | 629 | 51.47 | <0.001 | 252 | 54.78 |
| **CCU admission** | 18 | 0.75 | 60 | 3.57 | <0.001 | 12 | 0.98 | 0.460 | 48 | 10.43 |
| **Mortality** | 79 | 3.28 | 155 | 9.22 | <0.001 | 42 | 3.44 | 0.803 | 113 | 24.57 |
| **DNACPR** | 300 | 12.45 | 704 | 41.85 | <0.001 | 449 | 36.74 | <0.001 | 255 | 55.43 |
| **LOS (days)** |  |  |  |  |  |  |  |  |  |  |
| Median (IQR) | 1.42 (0.22-6.78) |  | 2.65 (0.51-6.86) |  | <0.001 | 1.82 (0.29-5.21) |  | 0.56 | 5.37 (2.11-11.05) |  |

The April 2020 cohort were more likely to be admitted to CCU (2020; 3.57% Vs. 2019; 0.75%, p< 0.001) and were more likely to die during their admission (mortality 2020; 9.22% Vs. 2019; 3.28%, p< 0.001). When COVID-19 admissions were excluded, there were no significant differences in either CCU admissions or mortality between the same months in the two years (Table 1).

The median length of stay was significantly longer in April 2020 than April 2019 (median (IQR) 2.65 days (0.51-6.86) vs 1.42 days (0.22-6.78), p< 0.001). When only non-COVID related admissions were studied, there was no difference in length of stay in April 2020 compared to April 2019.

Twenty-eight percent of medical admissions in April 2020 were due to COVID-19. Excluding those COVID-19 patients, admissions in 2020 had a different distribution of presentations compared to 2019 (Figure 2).

**Figure 2:**
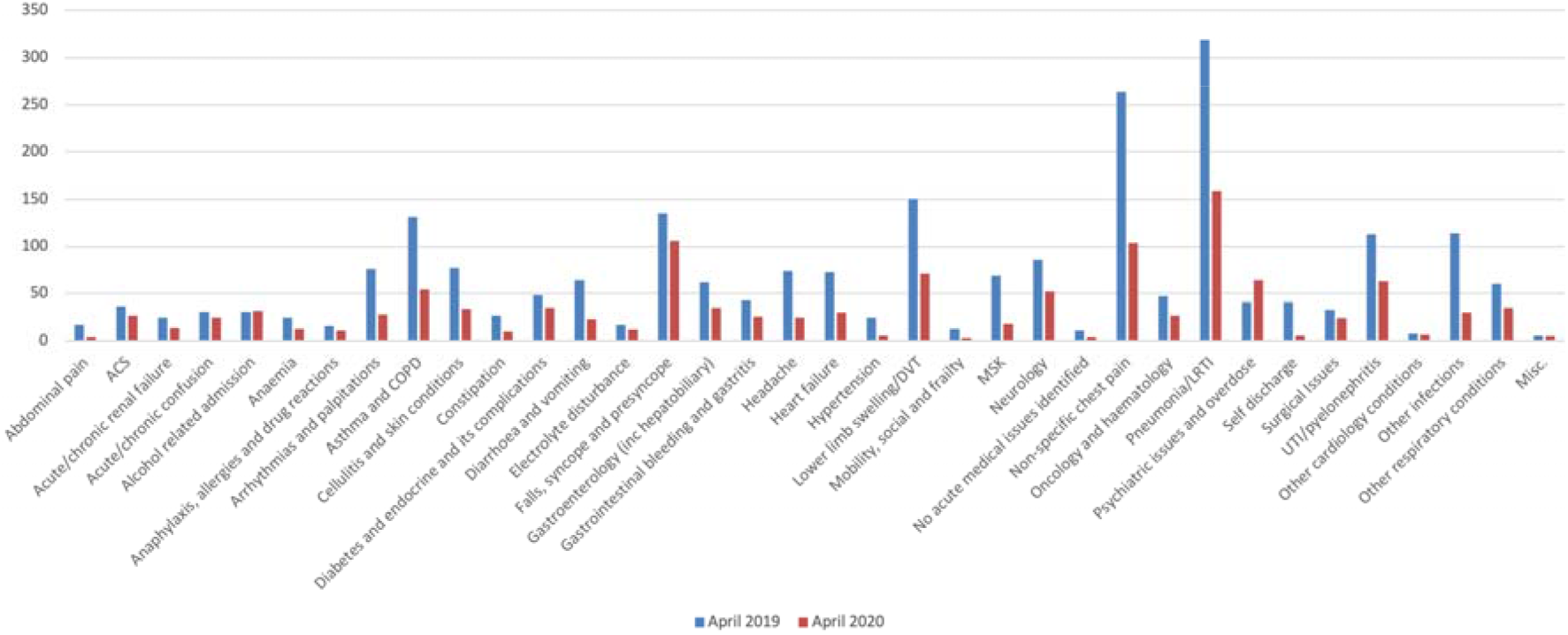
Discharge diagnosis for medical patients admitted in April 2019 and April 2020 (excluding patients with COVID-19). **Legend.** Diagnosis taken from discharge documentation for patients discharged from hospital, or from bereavement documentation for patients who died during admission. ACS = acute coronary syndrome; COPD = chronic obstructive pulmonary disease; DVT = deep vein thrombosis; LRTI = lower respiratory tract infection; MSK = musculoskeletal pain; UTI = urinary tract infection.

The absolute number of non-COVID admissions were reduced compared to the same period in 2019 for all diagnostic categories except alcohol-related admissions and psychiatric conditions and overdoses (Table 2). The number of admissions due to psychiatric conditions and overdoses rose in 2020.

**Table 2:**
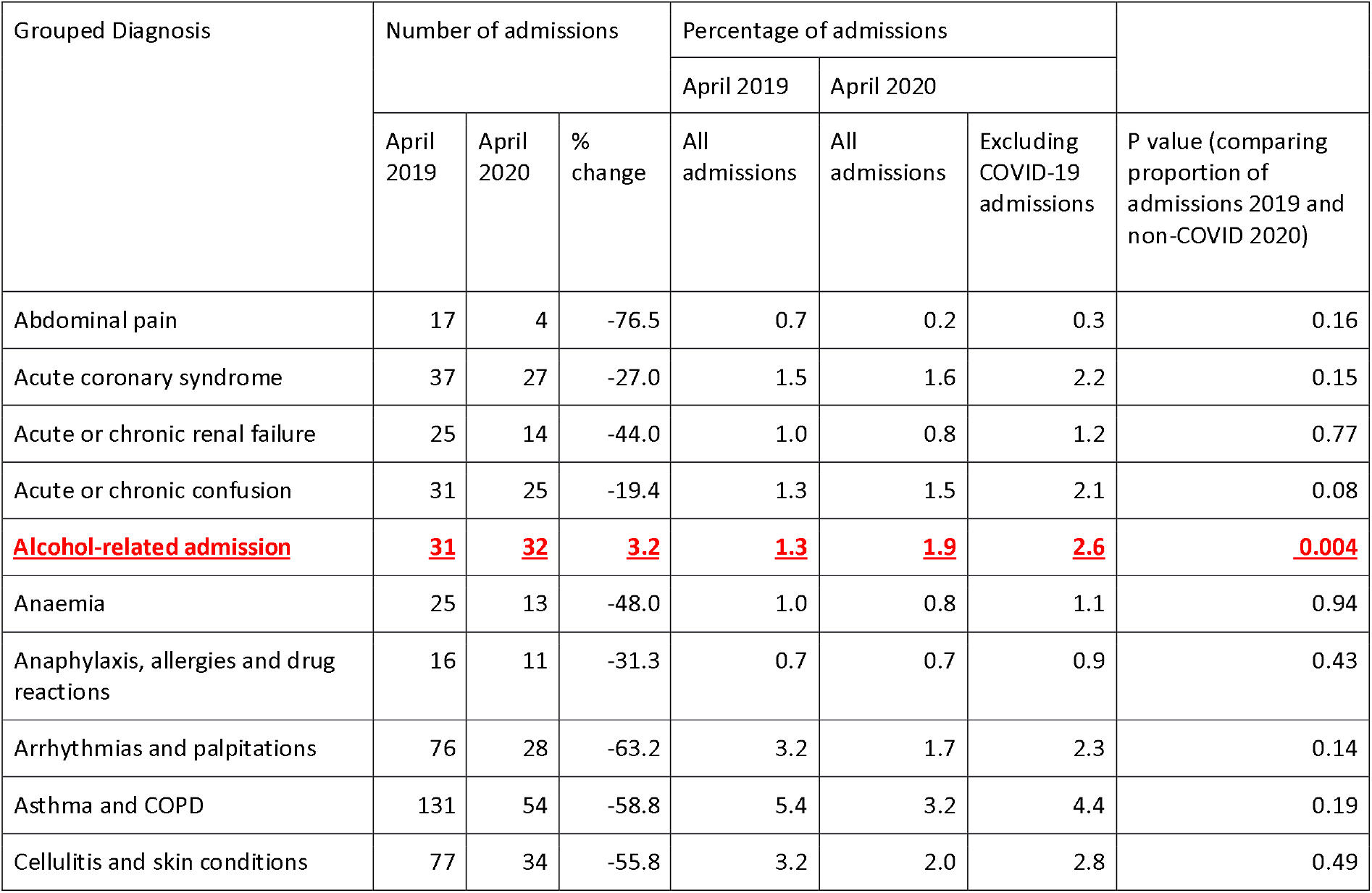
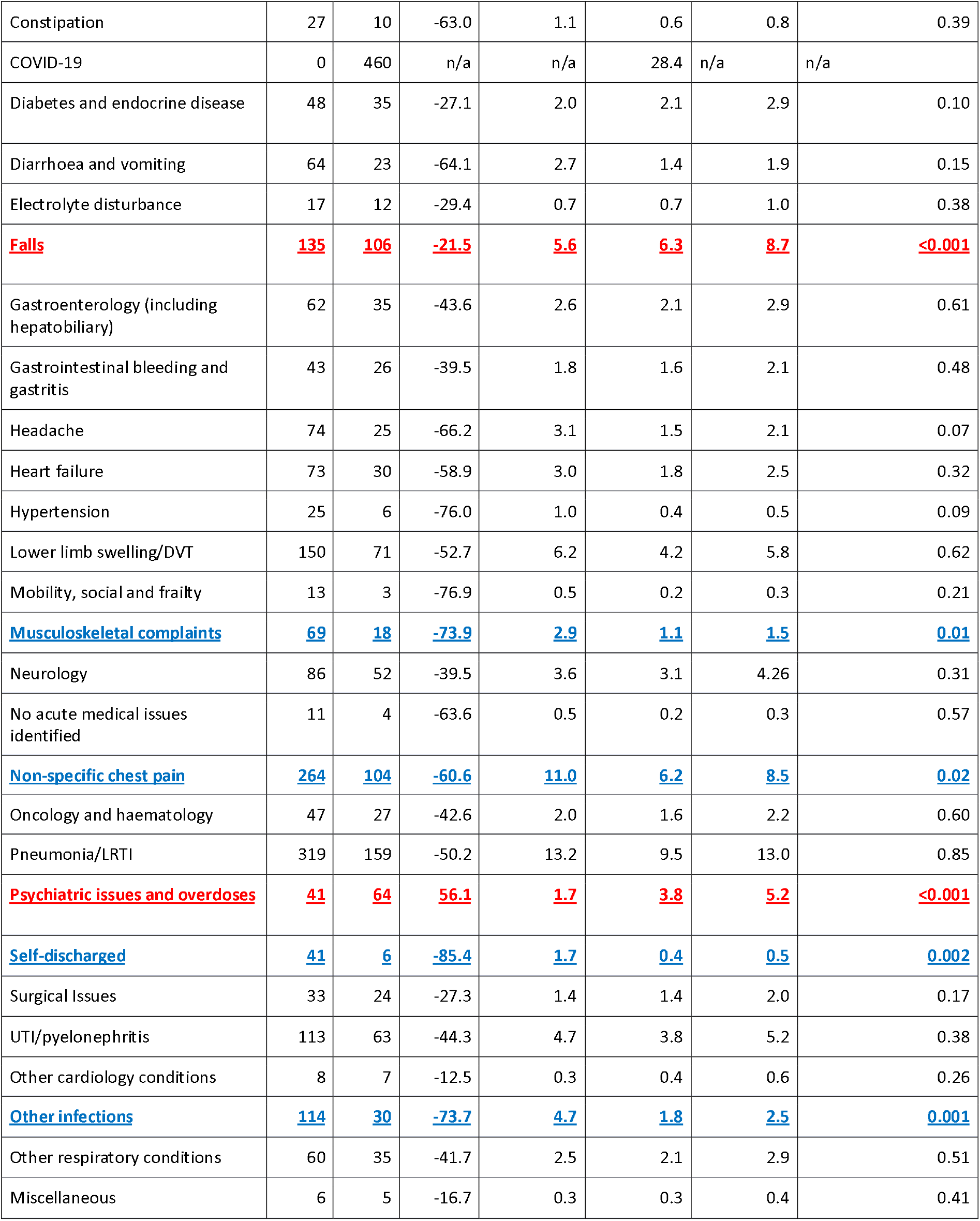
Discharge diagnoses for patients admitted as emergency medical admission. **Legend.** p-values are those when the proportion of admissions from April 2019 was compared to non-COVID admissions April 2020. COPD = chronic obstructive pulmonary disease; DVT = deep vein thrombosis; LRTI = lower respiratory tract infection; UTI = urinary tract infection. Blue text indicates where there was a significant decrease in the proportion of cases between the two months and red indicates where there was a significant increase in the proportion of cases between April 2019 and April 2020.

| Grouped Diagnosis | Number of admissions |  |  | Percentage of admissions |  |  | P value (comparing proportion of admissions 2019 and non-COVID 2020) |
| --- | --- | --- | --- | --- | --- | --- | --- |
|  |  |  |  | April 2019 | April 2020 |  |  |
|  | April 2019 | April 2020 | % change | All admissions | All admissions | Excluding COVID-19 admissions |  |
| Abdominal pain | 17 | 4 | -76.5 | 0.7 | 0.2 | 0.3 | 0.16 |
| Acute coronary syndrome | 37 | 27 | -27.0 | 1.5 | 1.6 | 2.2 | 0.15 |
| Acute or chronic renal failure | 25 | 14 | -44.0 | 1.0 | 0.8 | 1.2 | 0.77 |
| Acute or chronic confusion | 31 | 25 | -19.4 | 1.3 | 1.5 | 2.1 | 0.08 |
| <u>Alcohol-related admission</u> | <u>31</u> | <u>32</u> | <u>3.2</u> | <u>1.3</u> | <u>1.9</u> | <u>2.6</u> | <u>0.004</u> |
| Anaemia | 25 | 13 | -48.0 | 1.0 | 0.8 | 1.1 | 0.94 |
| Anaphylaxis, allergies and drug reactions | 16 | 11 | -31.3 | 0.7 | 0.7 | 0.9 | 0.43 |
| Arrhythmias and palpitations | 76 | 28 | -63.2 | 3.2 | 1.7 | 2.3 | 0.14 |
| Asthma and COPD | 131 | 54 | -58.8 | 5.4 | 3.2 | 4.4 | 0.19 |
| Cellulitis and skin conditions | 77 | 34 | -55.8 | 3.2 | 2.0 | 2.8 | 0.49 |
| Constipation | 27 | 10 | -63.0 | 1.1 | 0.6 | 0.8 | 0.39 |
| COVID-19 | 0 | 460 | n/a | n/a | 28.4 | n/a | n/a |
| Diabetes and endocrine disease | 48 | 35 | -27.1 | 2.0 | 2.1 | 2.9 | 0.10 |
| Diarrhoea and vomiting | 64 | 23 | -64.1 | 2.7 | 1.4 | 1.9 | 0.15 |
| Electrolyte disturbance | 17 | 12 | -29.4 | 0.7 | 0.7 | 1.0 | 0.38 |
| **Falls** | **135** | **106** | **-21.5** | **5.6** | **6.3** | **8.7** | **<0.001** |
| Gastroenterology (including hepatobiliary) | 62 | 35 | -43.6 | 2.6 | 2.1 | 2.9 | 0.61 |
| Gastrointestinal bleeding and gastritis | 43 | 26 | -39.5 | 1.8 | 1.6 | 2.1 | 0.48 |
| Headache | 74 | 25 | -66.2 | 3.1 | 1.5 | 2.1 | 0.07 |
| Heart failure | 73 | 30 | -58.9 | 3.0 | 1.8 | 2.5 | 0.32 |
| Hypertension | 25 | 6 | -76.0 | 1.0 | 0.4 | 0.5 | 0.09 |
| Lower limb swelling/DVT | 150 | 71 | -52.7 | 6.2 | 4.2 | 5.8 | 0.62 |
| Mobility, social and frailty | 13 | 3 | -76.9 | 0.5 | 0.2 | 0.3 | 0.21 |
| **Musculoskeletal complaints** | **69** | **18** | **-73.9** | **2.9** | **1.1** | **1.5** | **0.01** |
| Neurology | 86 | 52 | -39.5 | 3.6 | 3.1 | 4.26 | 0.31 |
| No acute medical issues identified | 11 | 4 | -63.6 | 0.5 | 0.2 | 0.3 | 0.57 |
| **Non-specific chest pain** | **264** | **104** | **-60.6** | **11.0** | **6.2** | **8.5** | **0.02** |
| Oncology and haematology | 47 | 27 | -42.6 | 2.0 | 1.6 | 2.2 | 0.60 |
| Pneumonia/LRTI | 319 | 159 | -50.2 | 13.2 | 9.5 | 13.0 | 0.85 |
| **Psychiatric issues and overdoses** | **41** | **64** | **56.1** | **1.7** | **3.8** | **5.2** | **<0.001** |
| **Self-discharged** | **41** | **6** | **-85.4** | **1.7** | **0.4** | **0.5** | **0.002** |
| Surgical issues | 33 | 24 | -27.3 | 1.4 | 1.4 | 2.0 | 0.17 |
| UTI/pyelonephritis | 113 | 63 | -44.3 | 4.7 | 3.8 | 5.2 | 0.38 |
| Other cardiology conditions | 8 | 7 | -12.5 | 0.3 | 0.4 | 0.6 | 0.26 |
| **Other infections** | **114** | **30** | **-73.7** | **4.7** | **1.8** | **2.5** | **0.001** |
| Other respiratory conditions | 60 | 35 | -41.7 | 2.5 | 2.1 | 2.9 | 0.51 |
| Miscellaneous | 6 | 5 | -16.7 | 0.3 | 0.3 | 0.4 | 0.41 |

When viewed as a proportion of admissions, alcohol-related admissions (p = 0.004), psychiatric conditions and overdoses (p< 0.001) comprised a larger proportion of total admissions in April 2020 than in April 2019. The proportion of patients who were in hospital due to falls, syncope or presyncope also increased in April 2020 (p< 0.001).

In April 2020 compared to April 2019, both the absolute number and the proportion of admissions that were due to non-specific chest pain, to musculoskeletal complaints and patients who self-discharged prior to assessment decreased (p = 0.02, p = 0.01 and p = 0.002 respectively).

The proportion of patients with a documented Do Not Attempt Cardiopulmonary Resuscitation (DNACPR) decision increased significantly between April 2019 and April 2020 for both COVID and non COVID admitting complaints and across all age groups (Table 3).

**Table 3:** DNACPR documentation rates based on demographics. **Legend.** Patients were divided into those with a ‘do not resuscitate’ order or those for full resuscitation in the event of a cardiac arrest for the two time periods. For 2020 data, patients were further divided into those without COVID-19. For age, p values compare those aged 16 – 55 years, 56 – 65 years, 66 – 75 years and over 76 year olds. Comparisons were made using a Fisher Exact test.

|  | 2019 |  | 2020 |  | P value<br>(all 2019<br>and 2020) | 2020 (Excluding COVID-19<br>admissions) |  | P value<br>(all 2019 and<br>non-COVID 2020) |
| --- | --- | --- | --- | --- | --- | --- | --- | --- |
|  | N | % | N | % |  | N | % |  |
| **Sex** |  |  |  |  |  |  |  |  |
| Male | 123/1054 | 11.7% | 339/881 | 38.5% | <0.0001 | 218/629 | 51.5% | <0.0001 |
| Female | 177/1355 | 13.1% | 365/801 | 45.6% | <0.0001 | 231/593 | 48.5% | <0.0001 |
| **Age** |  |  |  |  |  |  |  |  |
| 16-25 | 0/155 | 0.0% | 1/38 | 2.6% |  | 1/33 | 3.0% |  |
| 26-35 | 1/184 | 0.5% | 2/108 | 1.9% |  | 1/91 | 1.1% |  |
| 36-45 | 4/229 | 1.8% | 2/127 | 1.6% | <0.0001 | 2/93 | 2.2% | <0.0001 |
| 46-55 | 9/296 | 3.0% | 26/229 | 11.4% |  | 18/164 | 11.0% |  |
| 56-65 | 21/333 | 6.3% | 59/245 | 24.1% | <0.0001 | 38/179 | 21.2% | <0.0001 |
| 66-75 | 48/406 | 11.8% | 142/304 | 46.7% | <0.0001 | 93/227 | 41.0% | <0.0001 |
| 76-85 | 99/470 | 21.1% | 251/352 | 71.3% |  | 157/244 | 64.3% |  |
| 86-95 | 105/310 | 33.9% | 204/261 | 78.2% | <0.0001 | 129/180 | 71.7% | <0.0001 |
| 96+ | 13/26 | 50.0% | 17/18 | 94.4% |  | 10/11 | 90.9% |  |

## Discussion

This study compares acute medicine admissions over one month during the height of COVID-19 and the same month in 2019 and describes changes in health-seeking behaviour during these times.

Overall, the number of medical admissions fell in April 2020 compared to April 2019. The largest reduction in the proportion of admissions was for younger adults, with no significant change in the proportion of other age groups. Young adults access healthcare differently, with higher utilisation of urgent and emergency services^(6, 7)^. Further research should assess whether this reduction in hospital presentation was associated with any negative outcomes. If not, alternative, community health services may be more appropriate for some within this age group in the future.

The proportion of alcohol-related and psychiatric admissions increased. The COVID-19 pandemic may have impacted mental health for multiple reasons, including anxiety, social isolation and work and financial uncertainty^(8, 9)^. It is predicted this impact will be sustained and adequate services will need to be available for these patients^(10)^.

The proportion of admissions for non-specific chest pain and musculoskeletal complaints decreased. It is currently unclear if this reduced presentation was associated with potential harm. However, if this were not the case, alternative pathways avoiding hospital attendance may be possible.

The proportion of admissions that related to falls and syncope increased. It would be important to assess if social isolation contributed to this increased burden of falls, as has been described previously^(11)^.

Though the reasons for decreased medical admissions could not be explored, there may be several contributory factors. Admissions for pneumonia, asthma and COPD may have been reduced by public health measures leading to the reduction of other respiratory pathogens circulating in the community^(12)^. Public health messages may have discouraged people from attending hospital, particularly those with symptoms suggesting viral or lower respiratory tract infection, normally common reasons for attendance. Certain patient groups may be more concerned about attending hospital, for example the immunosuppressed. Services specifically introduced during the pandemic period bridging community and secondary care may have led to changes in patient attendance. The specific impact of these factors have not been evaluated here.

Overall, there was an increased proportion of patients admitted to CCU and a higher proportion of patients with a documented DNACPR decision across all diagnoses and age groups. These results are consistent with recent research from the same Trust^(13)^. Previous research suggests advanced care plans and decisions are under-recorded in acute medical admissions^(14)^. It will be of interest to determine if COVID-19 leads to a sustained change in medical practice. Of note, in those admitted for reasons other than COVID-19, there was no difference in in-hospital mortality, or in the proportion of patients admitted to critical care, despite the higher rate of documented DNACPR decisions.

This study has limitations. This is a single centre analysis. Patients were excluded if admitted directly to a specialty. These admission routes were available prior to April 2019, so will not affect comparison between years described here. However, centres without these admission routes may have a different alteration in the profile of admissions.

## Conclusion

It is recognized that non-COVID medical admissions decreased during the COVID-19 pandemic, but this short report highlights which conditions and patient groups were most affected. The greatest reductions were seen in young adults and for non-specific diagnoses which often do not require in-patient treatment (non-cardiac chest pain, for example). This suggests these admissions are avoidable, and potentially alternative services might be more appropriate. However, there was an increase in the proportion of medical admissions related to psychiatric conditions, alcohol and drug misuse and falls. This potentially highlights the risks of social isolation during lockdown and planning for future peaks of COVID-19 should ensure adequate support for people at risk of these conditions.

## Data Availability

 To facilitate knowledge in this area, the anonymised participant data and a data dictionary defining each field will be available to others through application to PIONEER via the corresponding author.  The data will be available upon request and following approval of a process to ensure ethical data governance and through a data access agreement.  Please contact the corresponding author for details.

## Author contributions

All authors contributed to the design of the study, analysis of the data, drafted the paper and reviewed and approved the final version of the manuscript.

## Acknowledgements

This work was supported by the PIONEER Acute Care Hub and HDR-UK Better Care Programme. Health Data Research UK is an initiative funded by UK Research and Innovation, Department of Health and Social Care (England) and the devolved administrations, and leading medical research charities. This work uses data provided by patients and collected by the NHS as part of their care and support. We would like to acknowledge the contribution of all staff, key workers, patients and the community who have supported our hospitals and the wider NHS at this time.

## References

1. Report of the Acute Medicine Task Force. Acute medical care. The right person, in the right setting - first time. https://www.acutemedicineorguk/wp-content/uploads/2014/04/RCP-Acute-Medicine-Task-Force-Reportpdf. 2007; DOA 8th August 2020.

2. Public Health England. Research and analysis. Syndromic surveillance: weekly summaries for 2020. https://www.govuk/government/publications/svndromic-surveillance-weeklv-summaries-for-2020. 2020;: DOA 1st September 2020.

3. Mafham MM, Spata E, Goldacre R, Gair D, Curnow P, Bray M, et al. COVID-19 pandemic and admission rates for and management of acute coronary syndromes in England. The Lancet. 2020;396(10248): 381–9.

4. Sapey E, Gallier S, Mainey C, Nightingale P, McNulty D, Corothers H, et al. Ethnicity and risk of death in patients hospitalised for COVID-19 infection in the UK: an observational cohort study in an urban catchment area. BMJ OPen Respiratory Research. 2020;doi: bmjresp-2020-000644:1-11.

5. World Health Organisation. WHO COVID-19 Case definition. WHO/2019-nCoV/Surveillance_Case_Definition/20201,. 2020; DOA 27th August 2020.

6. Gnani S, Morton S, Ramzan F, Davison M, Ladbrooke T, Majeed A, et al. Healthcare use among preschool children attending GP-led urgent care centres: a descriptive, observational study. BMJ open. 2016; 6(6): e010672–e.

7. McHale P, Wood S, Hughes K, Bellis MA, Demnitz U, Wyke S. Who uses emergency departments inappropriately and when - a national cross-sectional study using a monitoring data system. BMC Medicine. 2013; 11(1): 258.

8. Holmes EA, O’Connor RC, Perry VH, Tracey I, Wessely S, Arseneault L, et al. Multidisciplinary research priorities for the COVID-19 pandemic: a call for action for mental health science. The Lancet Psychiatry. 2020; 7(6): 547–60.

9. Brooks SK, Webster RK, Smith LE, Woodland L, Wessely S, Greenberg N, et al. The psychological impact of quarantine and how to reduce it: rapid review of the evidence. The Lancet. 2020;395(10227): 912–20.

10. Durcan G, O’Shea N, Allwood L. Covid-19 and the nation’s mental health. Forecasting needs and risks in the UK. https://www.centreformentalhealthorguk/sites/default/files/2020-05/CentreforMentalHealth_COVID_MH_Forecasting_May20pdf. 2020;: DOA 8th July 2020.

11. Hajek A, König H-H. The association of falls with loneliness and social exclusion: evidence from the DEAS German Ageing Survey. BMC geriatrics. 2017; 17(1): 204.

12. Tan JY, Conceicao EP, Sim XYJ, Wee LEI, Aung MK, Venkatachalam I. Public health measures during COVID-19 pandemic reduced hospital admissions for community respiratory viral infections. LID - S0195-6701(20)30354-6 [pii] LID - 10.1016/j.jhin.2020.07.023 [doi] FAU - Tan, J Y. J Hosp Infect. 2020;S0195–6701(20)30354-6(1532-2939 (Electronic)).

13. Coleman JJ, Botkai A, Marson EJ, Evison F, Atia J, Wang J, et al. Bringing into focus treatment limitation and DNACPR decisions: How COVID-19 has changed practice. Resuscitation. 2020; 155: 172–9.

14. Knight T, Malyon A, Fritz Z, Subbe C, Cooksley T, Holland M, et al. Advance care planning in patients referred to hospital for acute medical care: Results of a national day of care survey. EClinicalMedicine. 2020; 19.

